# Cerebral Small Vessel Disease Burden and Longitudinal Cognitive Decline from age 73 to 82: the Lothian Birth Cohort 1936

**DOI:** 10.1101/2021.03.28.21254499

**Authors:** OKL Hamilton, SR Cox, JA Okely, F Conte, L Ballerini, ME Bastin, J Corley, AM Taylor, D Page, AJ Gow, S Muñoz Maniega, P Redmond, M del C Valdés-Hernández, JM Wardlaw, IJ Deary

## Abstract

Slowed processing speed is considered a hallmark feature of cognitive decline in cerebral small vessel disease (SVD), however, it is unclear whether SVD’s association with slowed processing might be due to its association with overall declining general cognitive ability. We quantified the total MRI-visible SVD burden of 540 members of the Lothian Birth Cohort 1936 (age:72.6□0.7 years; 47% female). Using latent growth curve modelling, we tested associations between total SVD burden at mean age 73 and changes in general cognitive ability, processing speed, verbal memory, and visuospatial ability, measured at age 73, 76, 79 and 82. Covariates included age, sex, vascular risk, and childhood cognitive ability. In the fully-adjusted models, greater SVD burden was associated with greater declines in general cognitive ability (standardised β: -0.201; 95%CI: [-0.36, -0.04]; pFDR=0.022) and processing speed (−0.222; [-0.40, -0.04]; pFDR=0.022). SVD burden accounted for between 4 and 5% of variance in declines of general cognitive ability and processing speed. After accounting for the covariance between tests of processing speed and general cognitive ability, only SVD’s association with greater decline in general cognitive ability remained significant, prior to FDR correction (−0.222; [-0.39, -0.06]; p=0.008; pFDR=0.085). Our findings do not support the notion that SVD has a specific association with declining processing speed, independent of decline in general cognitive ability (which captures the variance shared across domains of cognitive ability). The association between SVD burden and declining general cognitive ability supports the notion of SVD as a diffuse, whole-brain disease and suggests that trials monitoring SVD-related cognitive changes should consider domain-specific changes in the context of overall, general cognitive decline.

## Introduction

Cerebral small vessel disease (SVD) is a major cause of cognitive impairment in older adults. It causes approximately 25% of all strokes and is the second most common cause of dementia after Alzheimer’s disease, either on its own or through mixed pathologies (1,2). Caused by dysfunction of the brain’s arterioles, capillaries, and venules, the downstream effects of SVD are visible on neuroimaging as white matter hyperintensities (WMH) and lacunes of presumed vascular origin, cerebral microbleeds, and visible perivascular spaces (PVS; see Figure 1) (3). In most individuals, these radiological markers do not result in overt clinical symptoms, however, their presence doubles the risk of stroke, and increases the risk of dementia and death in the general population (4). Despite its contribution to cognitive decline and to the development of associated co-morbidities (4,5), the precise nature of the associations between the radiological burden of SVD and decline in domain-specific cognitive abilities remains unclear.

**Figure 1:**
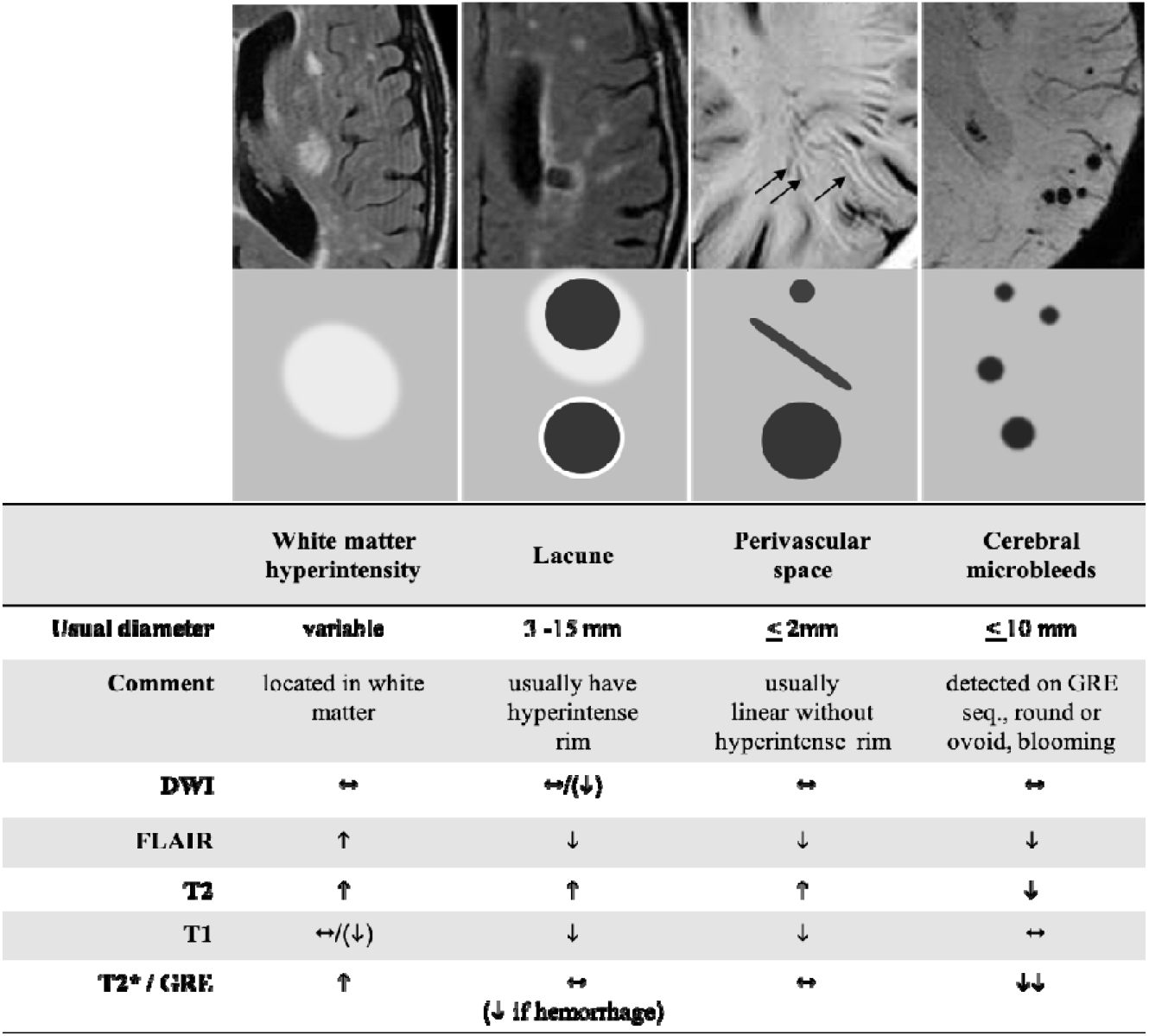
Key radiological markers of SVD examined in this study. *Figure 1 note:* Examples and schematic representations of key radiological features of SVD, according to STRIVE guidelines (3). Adapted with permission from Wardlaw et al. *The Lancet Neurology* 2013; 12(8): 822-38 [licence number 5010341055200, dated 15^th^ February 2021]. DWI: diffusion-weighted imaging; GRE: gradient-recalled echo.

Current consensus statements suggest that SVD is associated with declining processing speed and executive function, alongside relative preservation of memory and language abilities (6,7). However, previous studies examining domain-specific cognitive decline in SVD have not accounted for the well-replicated phenomenon in psychological research that cognitive test scores typically correlate positively with one another, such that an individual who performs well on a given cognitive test, is likely to perform well on a broader range of tests (8). This common variance among test scores can be accounted for by general cognitive ability, often termed ‘g’. General cognitive ability also accounts for the majority of variance in domain-specific cognitive decline; a recent meta-analysis estimated that, on average, 60% of the variance in cognitive changes were shared across abilities (9). It follows, therefore, that any domain-specific measure of cognitive ability will be influenced not only by an individual’s ability in that specific domain, but by their overall level of general cognitive ability. To be clear: if one finds an association between a biomarker, or any other exposure, and scores on a domain of cognitive ability or changes in a cognitive domain, there are three possible reasons for the result. First, the association might be wholly accounted for by an association with general cognitive ability (on which all cognitive domains load substantially); second, the association might be partly with general cognitive ability and partly with the cognitive domain; and, third, the association might indeed be exclusively with the cognitive domain. Thus, it is necessary to test formally whether previously-reported associations between SVD and processing speed are indeed specific to that domain, rather than confounded by the phenomenon of general cognitive ability.

To date, most studies investigating the relationship between SVD brain changes and cognitive decline have focused on individual radiological markers of SVD. WMH are the most frequently investigated SVD marker, perhaps due to their prevalence which is estimated at 64-94% in 82-year olds (10).

Recent meta-analyses report associations between greater baseline WMH burden and steeper decline in both general and domain-specific cognitive abilities, and greater risk of incident dementia (4,11). Similarly, the presence of lacunes and microbleeds have been associated with cognitive decline (12– 14), but associations between PVS and poorer cognitive ability, either cross-sectionally or longitudinally, are more variable (12). In recent years, several studies have quantified the ‘total brain burden’ of SVD using a simple 0-4 score, which allocates one point for the presence of each SVD marker (15–18). Whereas this approach goes some way towards considering the potential cumulative impact of different SVD markers on cognitive ability, the 0-4 score lacks sensitivity to subtle differences in the severity of the individual markers, and hence to their relative associations with cognitive abilities.

To improve the fidelity of SVD burden quantification, two previous studies (one using the same sample as the present study) utilised continuous neuroimaging variables to construct continuous SVD burden scores (19,20). In the first of these studies, Jokinen and colleagues (19) demonstrated associations between SVD burden (the average of standardised WMH, lacune, grey matter, and hippocampal volumes) and declining processing speed, executive function, memory, and general cognitive ability over a 3-year period. In a subsequent study from our own research group, using data from the LBC1936 (20), a continuous latent variable of SVD burden was negatively associated with latent variables of processing speed, verbal memory and visuospatial ability, in a structural equation modelling framework (SEM). However, after accounting for the shared variance between domain-specific scores (i.e. the variance attributable to general cognitive ability), only the association with processing speed remained. These findings suggest that the apparent associations we observed between SVD burden and domain-specific scores of verbal memory and visuospatial ability, were largely due to the confounding associations between SVD burden and general cognitive ability.

Longitudinal associations between SVD burden and declines in specific domains of cognitive ability, independent of their associations with general cognitive ability, have yet to be examined. Here, in a sample of relatively-healthy older individuals, we investigate associations between total MRI-visible SVD burden at age 73 and longitudinal cognitive decline between the ages of 73 and 82, a period that coincides with a substantial increase in dementia risk (21). Using growth curve modelling in a SEM framework, we separate the variance in cognitive test scores attributable to general cognitive ability from the variance attributable to the important, ageing-relevant domains of processing speed, memory, and visuospatial ability. This enables us to test whether SVD-related decline in specific cognitive domains are attributable to or independent of declining general cognitive ability.

## Methods

This study uses data from the Lothian Birth Cohort 1936 (LBC1936), a longitudinal study of cognitive, brain, and general ageing (22). In brief, the LBC1936 is an ongoing follow-up study to the Scottish Mental Survey 1947 (SMS1947; 23), which tested the cognitive abilities of 70,805 11-year-old children who were born in 1936 and were attending school in Scotland in 1947. Between 2004 and 2007, 1091 individuals, most of whom had taken part in the SMS1947, were recruited to the LBC1936. They have contributed to up to five waves of data collection at mean ages of about 70 (n=1091), 73 (n=866), 76 (n=697), 79 (n=550), and 82 (n=431) years. The present study includes data from Waves 2 to 5 of the LBC1936 (there was no MRI at Wave 1). Wave 2 MRI data were unusable for 51 of the 731 participants who underwent neuroimaging. Images belonging to a further 140 participants exhibited noise or motion artefacts that precluded the computational quantification of PVS, which due to the small size of the PVS (<3mm), is highly sensitive to such artefacts. Therefore, the remaining 540 participants constitute the baseline sample of this study (see Figure 2). Approvals for Waves 2 to 5 of the LBC1936 were obtained from the Scotland A Research Ethics Committee for Scotland (07/MRE00/58). All participants gave written, informed consent.

**Figure 2:**
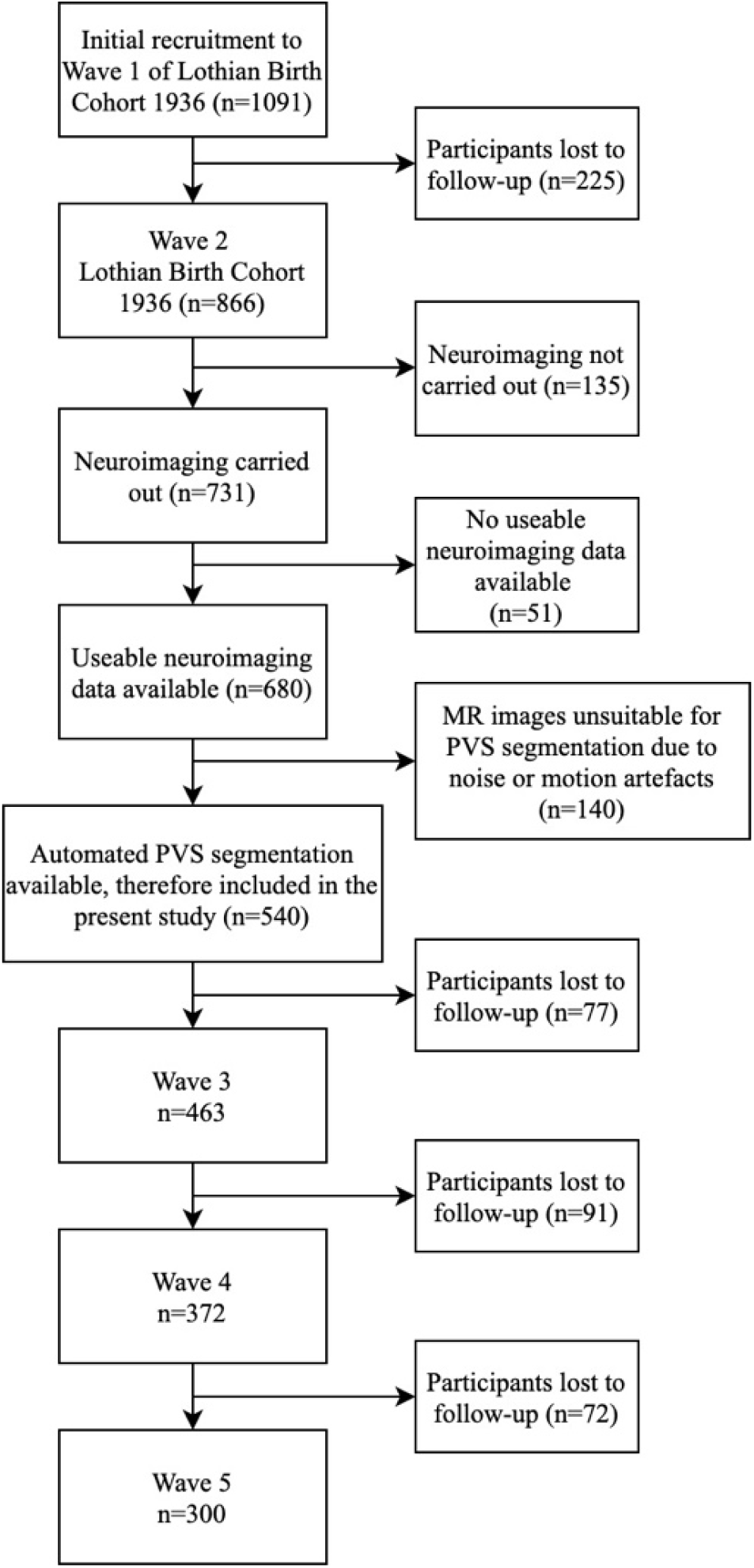
Consort diagram illustrating selection of the study sample.

### MRI data and the quantification of total SVD burden

The neuroimaging protocol for the LBC1936 has been published previously (24). Briefly, participants were scanned using a GE Signa Horizon HDx 1.5 Tesla clinical scanner (General Electric, Milwaukee, WI) operating in ‘research mode’, equipped with a self-shielding gradient set (33 mT/m maximum gradient strength) and manufacturer supplied eight-channel phased-array head coil.

Sequences acquired were T1-weighted (T1W), T2-weighted (T2W), T2*-weighted (T2*W) and fluid attenuated inversion recovery-weighted (FLAIR) images. Scanner stability was monitored throughout with a detailed quality assurance programme.

As previously reported, we used confirmatory factor analysis (CFA) to construct a latent variable representing total MRI-visible SVD burden (20). This latent variable comprised continuous WMH volume (divided by total intracranial volume (TIV) to account for differences in head size), continuous computationally-derived PVS count, and binary visual ratings of lacunes and microbleeds (i.e. present/absent). Total WMH volume and TIV, were measured semi-automatically using a validated multispectral image processing method that combines T2*W and FLAIR sequences in the colour space for enhanced feature discrimination in the segmentation (25), available from https://sourceforge.net/projects/bric1936/. All slices of all scans were checked by a trained observer and manually corrected, if necessary, to ensure that no true WMH had been omitted and to avoid including erroneous tissues in the WMH. PVS were computationally segmented in the native T2W space in the centrum semiovale using a recently validated technique (26,27) and are presented as total number of PVS. All binary PVS masks (superimposed on T2W images) were visually checked for the accurate quantification of PVS by a trained operator and were accepted or rejected blind to all other data. Where ambiguity arose, FLAIR and T1W sequences were also checked. Lacunes and microbleeds were rated by an experienced, registered neuroradiologist. Lacunes were classified as being present or absent and were defined as small (>3mm and <2cm in diameter) subcortical lesions of cerebrospinal fluid-equivalent signal on T2W and decreased signal on T1W and FLAIR images in the white matter, basal ganglia, and brainstem (3,24). Cerebral microbleeds were classified as being present or absent and were defined as small (<5mm), homogeneous, round foci of low signal intensity on T2*W images in the white matter, basal ganglia, brain stem, cerebellum, and cortico-subcortical junction (3,24). A random 20% sample of visual-ratings and any uncertain cases were independently checked by a second neuroradiologist, with disagreements resolved by consensus.

### Cognitive data

Participants completed the same series of cognitive tests at each wave of data collection, in the same location, administered using the same instructions. According to previous work examining their correlational structure (28), we grouped cognitive tests into the following domains:

1. Processing speed was measured using Digit Symbol Substitution and Symbol Search from the Wechsler Adult Intelligence Scale-III (WAIS-IIIUK; 29) and two experimental tasks: Inspection Time (30) and Four Choice Reaction Time (31). The Inspection Time task requires participants to select the longer of two vertical lines that are flashed on a computer screen for between 6 and 200 milliseconds. The measure used here was the number of correct responses out of a total of 150 trials. Four Choice Reaction Time scores were multiplied by -1 so that higher scores indicated better performance.
2. Memory consisted of Verbal Paired Associates (total score) and Logical Memory (total score) from the Wechsler Memory Scale III UK (WMS-IIIUK; 32), and Backward Digit Span (WAIS-IIIUK).
3. Visuospatial ability included Block Design and Matrix Reasoning (WAIS-IIIUK) and Spatial Span (average of forwards and backwards; WMS-IIIUK).

Our measure of general cognitive ability encompassed each of the above-mentioned tests. The Moray House Test Number 12 (MHT), a 71-item test of general cognitive ability, was also administered at the age of 11 as part of the Scottish Mental Survey 1947. In this study, we use the raw MHT score, which can range from 0-76 and subsequently refer to this variable as childhood cognitive ability.

### Covariates

We included age in years, sex, vascular risk, and childhood cognitive ability in all models, in a stepwise manner. Vascular risk variables included self-reported history of hypertension (yes/no); diabetes mellitus (yes/no); smoking status (ever/never); blood-derived glycated haemoglobin (% total HbA1c); blood-derived total cholesterol (mmol/l); and systolic and diastolic blood pressure (average of six readings: three seated and three standing), which were measured by trained nurses. We observed very little change in vascular risk variables over the four waves of testing (see Table 2), possibly as vascular risk factors such as hypertension and diabetes are relatively well managed in the LBC1936. Therefore, we considered baseline (Wave 2, age 73) vascular risk to be a sufficient representation of participants’ vascular status over the study period. We used CFA to construct a latent variable representing vascular risk as previously modelled in the LBC1936(33) and extracted its factor score for inclusion as a covariate. Childhood cognitive ability accounts for approximately half of the variance in later life cognitive ability (34). In part, this association might be mediated by increased SVD risk, which has also been found to associate with lower childhood cognitive ability (35,36). Therefore, as we expect childhood cognitive ability to attenuate the association between SVD burden and cognitive abilities measured at the age of 73 (Wave 2), we included MHT score measured at the age of 11 as a further covariate in our analyses. Time-invariant covariates (sex, baseline vascular risk, and childhood cognitive ability) were regressed on the outcomes of interest (the general and domain-specific cognitive intercepts and slopes) and were allowed to covary with one another and with the latent SVD burden variable. We specified time-variant covariates (age in years at each wave) as direct predictors of the observed cognitive and MRI variables.

**Table 1:** Characteristics of the study sample at each wave.

|  | n | Wave 2 (n=540) | n | Wave 3 (n=463) | n | Wave 4 (n=372) | n | Wave 5 (n=300) |
| --- | --- | --- | --- | --- | --- | --- | --- | --- |
| **Sociodemographic** |  |  |  |  |  |  |  |  |
| Age, years | 540 | 72.5 (0.5) | 463 | 76.2 (0.7) | 372 | 79.5 (0.6) | 300 | 82.0 (0.5) |
| Female, n (%) | 540 | 252 (46.7%) | 463 | 217 (46.9%) | 372 | 195 (52.4%) | 300 | 147 (49.0%) |
| Education, years | 540 | 10.9 (1.2) | 463 | 10.9 (1.2) | 372 | 10.9 (1.2) | 300 | 11.0 (1.2) |
| **Vascular risk** |  |  |  |  |  |  |  |  |
| Hypertension history, n (%) | 540 | 259 (48.0%) | 462 | 251 (54.3%) | 372 | 218 (58.6%) | 300 | 176 (58.7%) |
| Systolic blood pressure | 534 | 146.5 (18.0) | 458 | 147.4 (18.5) | 366 | 144.2 (17.9) | 294 | 147.2 (20.2) |
| Diastolic blood pressure | 534 | 79.7 (18.0) | 458 | 80.3 (9.8) | 366 | 78.2 (9.7) | 294 | 78.6 (10.5) |
| Diabetes history, n (%) | 540 | 54 (10.0%) | 463 | 52 (11.2%) | 371 | 42 (11.3%) | 300 | 33 (11.0%) |
| HbA1c mmol/mol | 518 | 39.0 (6.4) | 438 | 40.8 (7.1) | 348 | 40.4 (8.0) | 280 | 40.2 (8.0) |
| Cholesterol, mmol/l | 521 | 5.2 (1.1) | 426 | 5.0 (1.2) | 360 | 5.0 (1.2) | 284 | 4.9 (1.2) |
| Cardiovascular disease history, n (%) | 540 | 154 (28.5%) | 463 | 152 (32.8%) | 372 | 135 (36.3%) | 298 | 118 (39.6) |
| Smoking status, n (%) | 540 | Ever=274 (50.7%)<br>Never=266 (49.3%) | 462 | Ever=218 (47.2%)<br>Never=244 (52.8%) | 372 | Ever=204 (54.8%)<br>Never=168 (45.2%) | 300 | Ever=167 (55.7%)<br>Never=133 (44.3%) |
| **Cognitive** |  |  |  |  |  |  |  |  |
| Moray House Test age 11 (raw score, max 76) | 511 | 50.2 (11.9) | 437 | 50.9 (11.6) | 372 | 51.1 (11.7) | 283 | 51.2 (11.5) |
| **Neuroimaging** |  |  |  |  |  |  |  |  |
| WMH volume cm<sup>3</sup> | 537 | 12.22 (12.8) | 387 | 16.37 (15.3) | 309 | 20.49 (17.7) | 241 | 22.41 (18.8) |
| Total brain volume cm<sup>3</sup> | 533 | 993.66 (88.4) | 387 | 976.35 (88.5) | 309 | 965.41 (87.0) | 241 | 947.3 (85.5) |
| PVS count | 540 | 258.7 (94.6) |  |  |  |  |  |  |
| Lacunes, n (%) | 540 | Present=28 (5.1%)<br>Absent=512 (94.9%) |  |  |  |  |  |  |
| Microbleeds, n (%) | 540 | Present=65 (12.0%)<br>Absent=475 (88.0%) |  |  |  |  |  |  |

*Note for Table 1:* Values are mean (SD) unless otherwise specified. PVS: visible perivascular spaces; WMH: white matter hyperintensities of presumed vascular origin.

**Table 2:** Characteristics at each wave of study completers only.

|  | n | Wave 2 | n | Wave 3 | n | Wave 4 | n | Wave 5 |
| --- | --- | --- | --- | --- | --- | --- | --- | --- |
| **Sociodemographic** |  |  |  |  |  | 86.8 |  |  |
| Age, years | 300 | 72.5 (0.7) | 297 | 76.2 (0.7) | 296 | 79.3 (0.6) | 300 | 82.0 (0.5) |
| Female, n (%) | 300 | 147 (49.0%) | 300 | 145 (48.8%) | 296 | 143 (48.3%) | 300 | 147 (49.0%) |
| Education, years | 300 | 11.0 (1.2) | 297 | 11.0 (1.2) | 296 | 11.0 (1.2) | 300 | 11.0 (1.2) |
| **Vascular risk** |  |  |  |  |  |  |  |  |
| Hypertension history, n (%) | 300 | 135 (45.0%) | 297 | 157 (52.9%) | 296 | 172 (58.1%) | 300 | 176 (58.7%) |
| Systolic blood pressure | 297 | 145.6 (18.0) | 293 | 146.4 (17.5) | 291 | 144.2 (17.9) | 294 | 147.2 (20.2) |
| Diastolic blood pressure | 297 | 79.4 (9.2) | 293 | 80.0 (9.4) | 291 | 78.4 (9.5) | 294 | 78.6 (10.5) |
| Diabetes history, n (%) | 300 | 20 (6.7%) | 297 | 26 (8.8%) | 296 | 31 (10.5%) | 300 | 33 (11.0%) |
| HbA1c mmol/mol | 290 | 38.8 (5.8) | 283 | 40.7 (7.1) | 281 | 40.5 (7.9) | 280 | 40.2 (8.0) |
| Cholesterol, mmol/l | 291 | 5.3 (1.1) | 275 | 5.1 (1.2) | 287 | 5.1 (1.1) | 284 | 4.9 (1.2) |
| Cardiovascular disease history, n (%) | 300 | 83 (27.7%) | 297 | 103 (34.7) | 296 | 110 (37.2%) | 298 | 118 (39.6) |
| Smoking status, n (%) | 300 | Ever=141 (47.0%)<br>Never=159 (55.0%) | 297 | Ever=134 (45.1%)<br>Never=163 (54.9%) | 296 | Ever=137 (46.3%)<br>Never=159 (53.7%) | 300 | Ever=133 (44.3%)<br>Never=167 (55.7%) |
| **Cognitive** |  |  |  |  |  |  |  |  |
| Moray House Test age 11 (max 76) | 283 | 51.2 (11.5) | 280 | 51.2 (11.5) | 279 | 51.3 (11.5) | 283 | 51.2 (11.5) |
| **Neuroimaging** |  |  |  |  |  |  |  |  |
| Total WMH volume cm<sup>3</sup> | 298 | 10.47 (11.3) | 258 | 14.76 (14.6) | 252 | 19.17 (16.9) | 241 | 22.41 (18.8) |
| Total brain volume cm<sup>3</sup> | 300 | 991.9 (87.2) | 258 | 976.0 (86.8) | 252 | 964.4 (88.0) | 241 | 947.3 (85.5) |
| PVS count | 300 | 251.8 (92.5) |  |  |  |  |  |  |
| Lacunes, n (%) | 300 | Present=13 (4.3%)<br>Absent=287 (95.7%) |  |  |  |  |  |  |
| Microbleeds, n (%) | 300 | Present=33 (11.0%)<br>Absent=267 (89.0%) |  |  |  |  |  |  |

*Note for Table 2:* Values are mean (standard deviation) unless otherwise specified. PVS: visible perivascular spaces; WMH: white matter hyperintensities of presumed vascular origin. Three participants did not take part in Wave 3 only and four participants did not take part in Wave 4 only.

### Statistical Analysis

#### Measurement models

First, we used hierarchical ‘factor-of-curves’ models (FoC) within a SEM framework (37), as has been done previously in this cohort (38,39), to examine the initial level and subsequent decline in general cognitive ability, processing speed, memory, and visuospatial ability between mean ages 73 (Wave 2), 76 (Wave 3), 79 (Wave 4), and 82 (Wave 5). A FoC model estimates the initial level of each cognitive test (intercept) and its trajectory across all four waves of testing (slope). The latent intercepts and slopes of each cognitive test load onto superordinate latent intercepts and latent slopes of their respective cognitive domains (see Figure 3A). This permits analysis of the initial level and trajectory of each cognitive domain as if they were directly observed.

**Figure 3:**
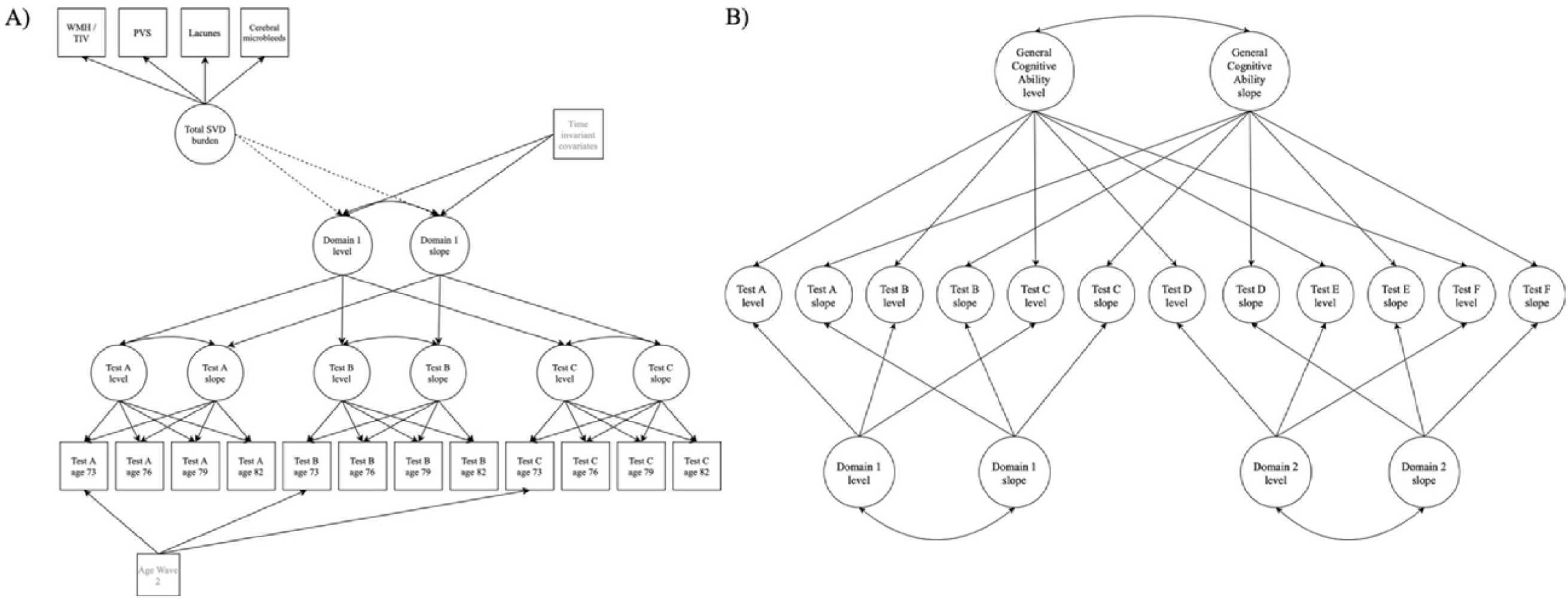
Illustrations of a hierarchical ‘factor-of-curves’ model and a longitudinal bifactor model of cognitive ability. *Note for Figure 3A:* A hierarchical ‘Factor-of-curves’ (FoC) model of cognitive ability. For the hierarchical FoC models, a growth curve was estimated for each individual cognitive test, producing a latent intercept and slope. These test-specific latent intercepts and slopes in turn loaded onto an overall latent intercept and slope for the cognitive domain. Loadings on the slopes were set to 0, 3.78, 6.83, and 9.55, to reflect the average time lags between baseline and subsequent waves. In this illustration we also show how we specified associations between latent SVD burden and the intercept and slope of the cognitive factor (see dashed lines), and how we included additional time-invariant (sex, vascular risk, childhood cognitive ability) and time variant (age) covariates (see items in grey). Separate models were carried out for each cognitive domain (i.e. general cognitive ability, processing speed, verbal memory, and visuospatial ability). Following conventional SEM notation, variables in squares were observed and measured, and variables in circles represent unobserved latent variables. Single headed arrows represent specified relationships between variables and double headed arrows represent correlations. *Note for Figure 3B:* Longitudinal bifactor model. In the centre of the model are the latent intercept and slope of each cognitive test, which were constructed using latent growth curves of the originally observed test scores at each time point (as described in the panel A note). The variance in these test-specific latent intercepts and slopes is separated into that which contributes to the latent intercept and slope of each cognitive domain, and that which contributes to the latent intercept and slope of general cognitive ability. We tested associations between total SVD burden and the intercept and slope of each cognitive variable simultaneously (not shown in this illustration). Additional time-invariant and time-variant covariates were included as indicated for the hierarchical FoC model (not shown in this illustration, see panel A for details).

#### Hierarchical factor-of-curves models of SVD-cognitive ability associations

Next, we specified linear regressions between baseline SVD burden (independent variable) and the latent intercepts and slopes of general cognitive ability, processing speed, memory, and visuospatial ability (dependent variables). This was carried out in separate hierarchical FoC models: one for general cognitive ability, and one for each cognitive domain. Importantly, it should be noted that at this stage the cognitive domains will contain any variance actually attributable to general cognitive ability (i.e. common across cognitive domains). Associations between SVD burden and latent cognitive intercepts approximate the cross-sectional associations that we reported previously (20).

Therefore, in this study we are primarily interested in the associations between SVD burden and the latent cognitive slopes.

#### Longitudinal bifactor models of SVD-cognitive ability associations: accounting for covariance between domain-specific scores and general cognitive ability

Previous analyses in the LBC1936 have estimated that general cognitive ability explains approximately 50% of the variability in the decline of individual test scores between the ages of 70 and 76, and up to 70% of variability in their decline between 70 and 79(38,40). Owing to this shared variability, estimations of decline in any measure of domain-specific cognitive ability will contain both the amount of decline in that domain-specific ability and the amount of decline in general cognitive ability. Therefore, to assess decline in domain-specific cognitive abilities, the variance in test scores associated with general cognitive ability must be removed. To do this, we constructed a longitudinal bifactor model (in a SEM framework) in which the variance associated with general and domain-specific abilities is parsed into separate latent variables, so that the level and trajectory of general cognitive ability and those of the domain-specific abilities can be measured independently of one another (see Figure 3B). To test associations between SVD burden and the level and decline of domain specific-abilities independently of general ability, we specified linear regressions between SVD burden (independent variable) and the latent intercepts and slopes of the cognitive variables (general cognitive ability and the three cognitive domains) from the longitudinal bifactor model.

#### Examining the contribution of WMH to the total SVD burden-cognitive ability associations

Of the radiological markers of SVD examined here, WMH are the most regularly associated with poorer cognitive abilities. It is plausible, therefore, that any associations between the SVD burden variable and cognitive factors could largely be driven WMH. To test whether this was the case, we repeated the hierarchical FoC analyses with WMH/TIV in the place of SVD burden as the predictor variable. As an indication of the relative utility of total SVD burden and WMH/TIV as predictors of cognitive outcomes, we examined the magnitude of standardised effect sizes and confidence intervals of models specifying WMH/TIV, and those of models specifying total SVD burden as the predictor.

As absolute fit indices are unavailable when using maximum likelihood to estimate models including binary measures, we assessed the fit of models specifying total SVD burden as the predictor (binary measures of lacunes and microbleeds contribute to the latent SVD variable) against a less-restrictive neighbouring model (i.e. one in which latent SVD burden, latent cognitive intercepts and latent cognitive slopes were permitted to correlate with one another) using the likelihood ratio test statistic calculated as follows: -2 x (loglikelihood of the less-restrictive model – loglikelihood of full model). For models specifying WMH/TIV as the predictor, model fit was assessed using four absolute fit indices: Root Mean Square Error of Approximation (RMSEA; <0.06 considered acceptable), Comparative Fit Index (CFI; >0.95 acceptable), Tucker-Lewis Index (TLI; >0.95 acceptable), and Standardized Root Mean Square Residual (SRMR; <0.08 acceptable; 41).

All analyses were carried out in MPlus version 8.4 (42) and were estimated using Full Information Maximum Likelihood (FIML), which estimates model parameters based on all available data from our sample of 540 LBC1936 participants. We corrected p-values for multiple comparisons using the False Discovery Rate adjustment (FDR; 43), with p.adjust in R version 4.0.1(44). This correction was carried out separately for p values from three different groups of models: 1) associations between total SVD burden and the intercept and slopes of cognitive change from hierarchical FoC models; 2) associations between WMH/TIV and the intercept and slopes of cognitive change from hierarchical FoC models; 3) associations between total SVD burden and the intercept and slopes of cognitive change from bifactor models.

## Results

### Cohort characteristics

Sociodemographic and clinical characteristics of participants at each wave are presented in Table 1. The same characteristics at each wave for study completers only are shown in Table 2, highlighting changes in the same sample over the four waves. We included 540 participants at baseline and lost between 13 and 16% of participants to follow-up at each subsequent wave. Participants lost to follow-up had a higher baseline prevalence of diabetes (but slightly lower cholesterol levels), greater total WMH volume, fewer years of education, and slightly lower childhood cognitive ability than study completers (see Table S1).

### Cognitive decline effect sizes between age 73 and 82

We first modelled the mean change in general cognitive ability, processing speed, memory, and visuospatial ability between the ages of 73 and 82 in separate hierarchical FoC models. Note that, at this stage, the cognitive domains will still contain any variance due to general cognitive ability. Table S2 provides details of the initial levels (intercepts) and trajectories (slopes) for each cognitive domain. All cognitive domain scores showed a significant mean decline over the nine-year period (all p<0.001). In standard deviation units, the declines per year were: -0.13 (just under 2 IQ points, which each have a SD of 15) for general cognitive ability, -0.16 for processing speed, -0.005 for memory, and 0.08 for visuospatial ability.

### Total SVD burden associations with declines in general cognitive ability and cognitive domains in separate factor-of-curves models

The key analyses in this study were associations between total SVD burden and trajectories of general cognitive ability, processing speed, verbal memory, and visuospatial ability. Note that in the hierarchical FoC models, the domains still contain any variance due to general cognitive ability. After the inclusion of covariates, total SVD burden was negatively associated with the slope of general cognitive ability (standardised β: -0.201; 95%CI: [-0.36, -0.04]; p=0.015; pFDR=0.022) and processing speed (−0.222; [-0.40, -0.04]; p=0.015; pFDR=0.022), but not with verbal memory or visuospatial ability (see Table 3). R^2^ values indicated that total SVD burden accounted for approximately 4% of the variance in the slope of general cognitive ability, and 5% of the variance in the slope of processing speed (which still contains general cognitive ability variance). In line with our previously reported results (20), total SVD burden was negatively associated with the intercept of all cognitive variables after the inclusion of covariates: (standardised betas ranged between -0.322 to - 0.173; pFDR ≤ 0.022; for full results see Table S3).

**Table 3:**
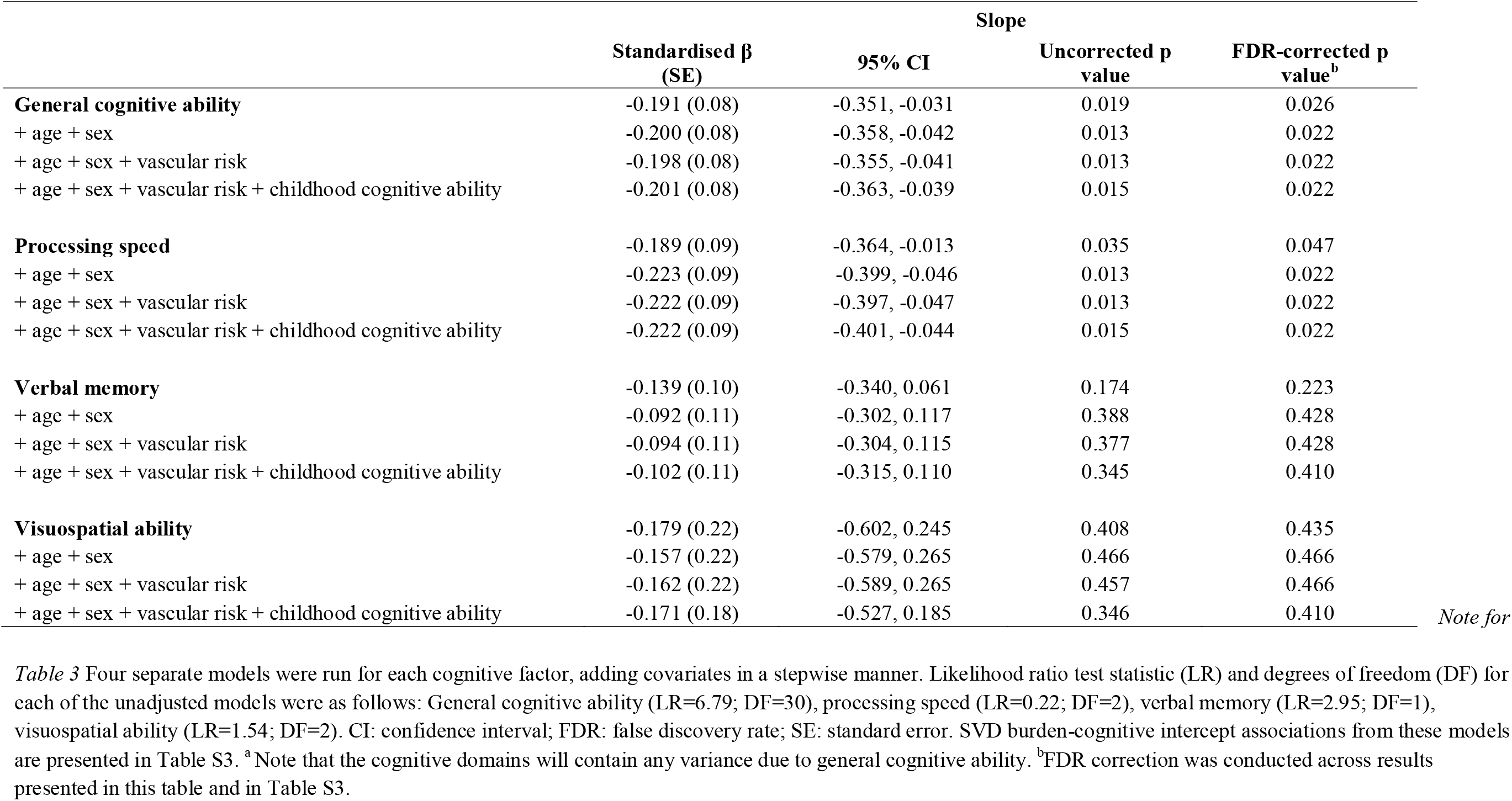
Factor-of-curves models of associations between total SVD burden and the slope of latent cognitive variables between the ages of 73 and 82^a^

### Total SVD burden associations with declines in general cognitive ability and specific cognitive domains, modelled simultaneously in a longitudinal bifactor model

We next tested associations between total SVD burden and cognitive variables using a bifactor model, which separates out the variance in cognitive test scores attributable to general cognitive ability and to domain-specific factors (see Table 4). Results of the fully-adjusted bifactor model indicated that total SVD burden was associated with greater decline (steeper downward slope) in general cognitive ability only prior to FDR correction (standardised β: -0.222; 95%CI: [-0.39, -0.06]; p=0.008; pFDR=0.085). We found no significant associations between total SVD burden and the slopes of any other cognitive variables (i.e. processing speed, verbal memory or visuospatial ability) in the bifactor model. In terms of SVD-cognitive intercept associations, total SVD burden was associated with the intercept of general cognitive ability only, but this association became non-significant after the inclusion of covariates and adjustment for FDR (see Table S4).

**Table 4:**
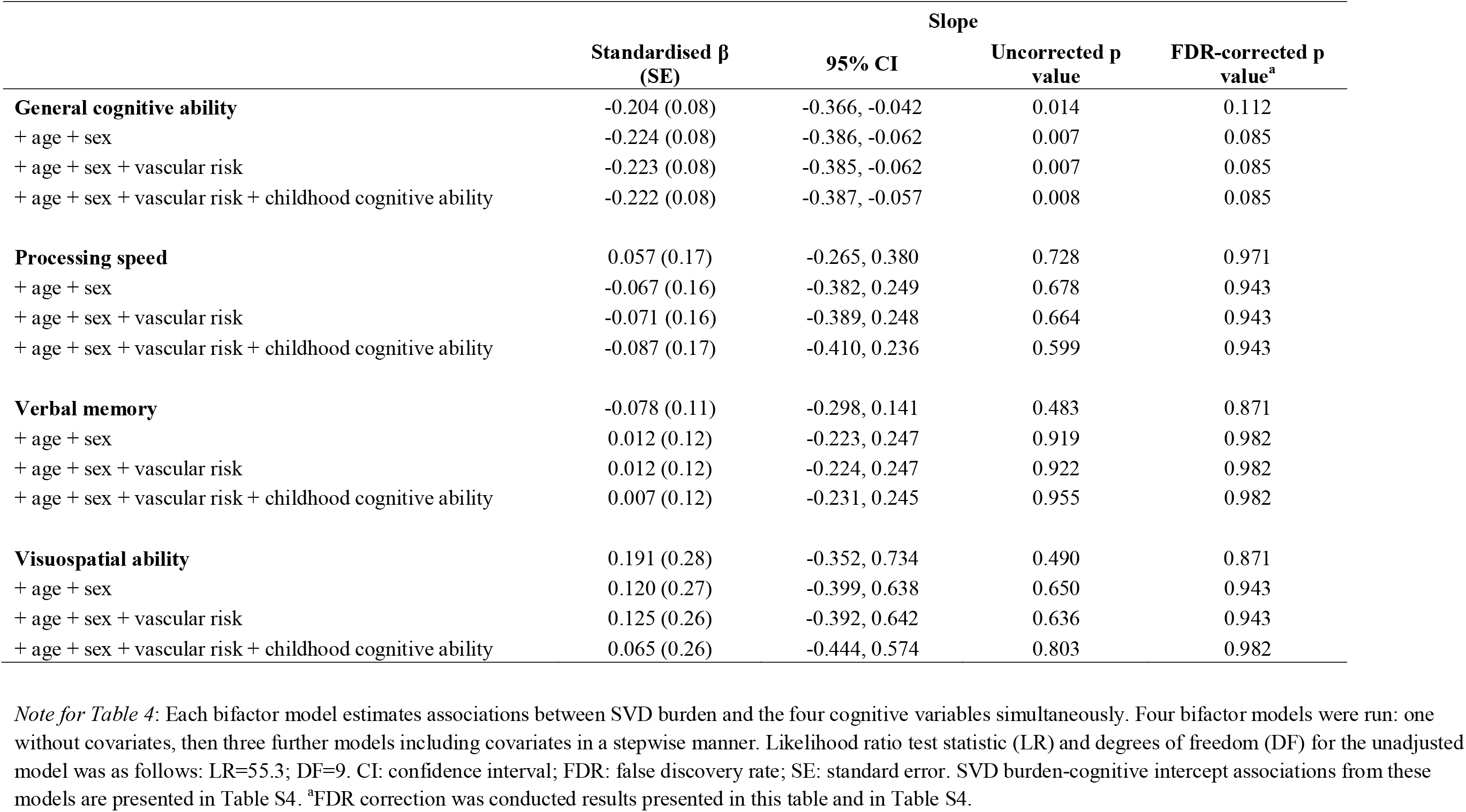
Results of bifactor models of associations between total SVD burden and slope of latent cognitive variables between age 73 and 82

Finally, we tested whether the associations observed between SVD burden and cognitive decline were likely driven by the contribution of WMH to the SVD burden score. We re-ran the hierarchical FoC models with WMH/TIV in the place of total SVD burden as the predictor (for results see Tables S5 and S6). Note that in the hierarchical FoC models, the domains will still contain any variance due to general cognitive ability. In the fully-adjusted, FDR-corrected models, we observed significant associations between WMH/TIV and the slopes of general cognitive ability (standardised β: -0.149; 95%CI: [-0.26, -0.04]; p=0.008; pFDR=0.012) and processing speed (standardised β: -0.176; 95%CI: [-0.30, -0.05]; p=0.007; pFDR=0.012). Effect sizes of these models were 0.021 and 0.027 standard deviations smaller (for general cognitive ability and processing speed, respectively) than models specifying total SVD burden as the predictor, and confidence intervals from models using different predictors overlapped substantially. Differences in effect size magnitudes were more pronounced for the total SVD-cognitive intercept associations; effect sizes of models with WMH/TIV as the predictor were between 0.06 and 0.12 standard deviations smaller than models with total SVD burden as the predictor. Overlap between confidence intervals of these models was present but more modest than for slopes.

## Discussion

In this longitudinal study of 540 community-dwelling older adults, we investigated associations between the total MRI-visible burden of cerebral SVD and the nine-year trajectory of cognitive abilities between the ages of 73 and 82. We found associations between greater SVD burden and greater decline in both general cognitive ability and processing speed, after accounting for age, sex, vascular risk and childhood cognitive ability. We then separated the variance in cognitive test scores attributable to domain-specific abilities and to general cognitive ability (using a bifactor model), to test SVD’s relationship with declining processing speed, independent of the influence of general cognitive decline. In the fully-adjusted bifactor model, the association between greater SVD burden and declining general cognitive ability was nominally significant (p=0.008), but became non-significant after FDR correction (pFDR=0.085). In contrast, in the bifactor model the negative association between total SVD burden and declining processing speed was non-significant both prior to and following FDR correction (p=0.599; pFDR=0.943). We were cautious in our use of FDR correction; smaller p-values in the bifactor models were heavily penalised due to the large number of p-values included in the correction. In addition to the non-zero-containing confidence intervals for this association, the overall results from this bifactor model suggest that SVD burden’s association with declining processing speed might be accounted for by overall decline in general cognitive ability. By overlooking the shared variance among domain-specific cognitive tests, previously observed associations between radiological markers of SVD and decline in domain-specific abilities could be an artefact of the relationship between SVD markers and declining general cognitive ability.

Alongside these main results, in the hierarchical FoC models we observed associations between total SVD burden and the initial levels of general cognitive ability, processing speed, verbal memory, and visuospatial ability at the age of 73. These results are in line with our previous analyses that used only cross-sectional data from age 73 (20).

Effect sizes for associations between total SVD burden and greater decline in general cognitive ability and processing speed (before accounting for their covariance), were medium sized (standardised betas were -0.201 and -0.222 respectively) (45). These relatively modest effect sizes are unsurprising considering the huge number of additional structural brain variables, such as decreasing white matter microstructural integrity and cortical volumes, that contribute to cognitive decline in later life (39,46). The addition of age, sex, vascular risk, and childhood cognitive ability did not attenuate these effect sizes. The lack of attenuation is expected as it has been observed previously in the LBC1936 that childhood cognitive ability is strongly associated with levels of cognitive ability in later life, but not with cognitive decline (38). Additionally, combined vascular risk factors measured in later life only account for approximately 2% of the variance in WMH in the LBC1936 (47; see also 48), and individually, factors such as a diagnosis of hypertension, diabetes, or cerebrovascular disease have shown no unique association with cognitive decline in this sample (28,38).

That SVD burden appears to be associated with an overall decline in general cognitive ability (after accounting for covariance between general and domain-specific test scores) is consistent with the pattern of non-pathological, age-related cognitive decline. Previous studies have estimated that the majority of the variance in age-related decline across domain-specific cognitive abilities is shared, and that the proportion of shared variance increases with age (up to 70% by the age of 85) (9,38). This implies that to a large and increasing extent, different domains of cognitive ability will decline together with advancing age. As age is the most important risk factor for SVD, it follows that SVD-related decline in domain-specific cognitive abilities are likely attributable to cognitive decline more generally. When examining associations between SVD burden and cognitive decline, our results suggested that SVD burden was associated with decline in general cognitive ability and processing speed only. However, it would be inaccurate to conclude from this that SVD-related cognitive decline does not involve declining visuospatial and memory abilities, as variance associated with decline in memory and visuospatial tests is well represented in the latent slope of general cognitive ability.

The potential association between SVD burden and overall decline in general cognitive ability also supports the notion of SVD as a diffuse, whole-brain disease that disrupts or ‘disconnects’ regions of the brain that sub-serve our cognitive abilities (49,50). Diffusion imaging (dMRI), which quantifies the diffusion of water molecules, thus providing a measurement of the microstructural organisation of the brain’s white matter, has demonstrated that SVD-related structural changes extend beyond visible radiological markers of the disease into the ‘normal appearing’ tissue that surrounds the visible lesion (51–53). Radiological markers of SVD are also known to have deleterious effects on areas remote from the lesion site; lacunes have been associated with thinning of the overlying cortical area, possibly due to degradation of the connecting white matter fibres (54). Widespread alterations of white matter connections have been associated with poorer cognitive abilities directly (55,56), and have also been highlighted by studies applying graph theoretic approaches to dMRI tractography data as a potential determinant of cognitive impairments via reduced density of white matter connections and impaired efficiency of information transfer between different brain regions (57–59).

To test whether associations between total MRI-visible SVD burden and cognitive outcomes were driven primarily by the contribution of WMH burden to the total SVD burden variable, we re-ran our hierarchical FoC models specifying WMH volume as the predictor. WMH volume was associated with the intercept of all cognitive factors, however, the magnitudes of effect sizes of these models were smaller (by between 0.06 and 0.12 standard deviations, with 95% Cis slightly overlapping) than those from models specifying SVD burden as the predictor. This suggests that total SVD burden could be a more powerful predictor of cognitive performance cross-sectionally than WMH burden alone. Interestingly, this was not the case when modelling cognitive decline; differences between the effect sizes of models specifying total SVD burden vs. WMH/TIV as the predictor of decline in cognitive outcomes appeared to be more modest (differences of between 0.021 and 0.027 standard deviations, with 95% Cis largely overlapping). Whereas incorporating measurements of PVS, lacunes and microbleeds alongside WMH in a single SVD burden score appears to strengthen the prediction of cognitive outcomes cross-sectionally, doing so may provide limited predictive power beyond that of WMH volume alone in associations with cognitive change between the ages of 73 and 82. WMH on neuroimaging represent heterogenous changes in the underlying brain tissue and cerebral microvasculature, ranging from alterations in water content and the build-up of perivascular oedema, which can resolve over time, to demyelination and axonal degeneration, which likely cannot (2). On the one hand, even though WMH are dynamic in nature (60), as one of the earliest radiological features of SVD, extensive WMH could indicate a longer duration of disease processes, thus could be more strongly related to detectable clinical features such as cognitive decline. On the other hand, other radiological markers of SVD such as lacunes and cerebral microbleeds, which represent more established vascular damage, are relatively uncommon in our study sample; only 28 participants from our sample of 540 had lacunes, and only 65 had microbleeds at baseline. Therefore, in a population of individuals with more severe SVD pathology, a variable representing total SVD burden may have more predictive power in relation to cognitive change. Additionally, the latent SVD burden variable represents only the shared variance between the four MRI markers of SVD. If the variance unique to each MRI marker of SVD also associates with cognitive change, the fact that it is not represented in our latent SVD burden variable may limit the magnitude of associations between the SVD burden variable and cognitive slopes.

This study benefits from the availability of multiple waves of in-depth cognitive testing in a relatively large sample of individuals, over almost a decade of time. In-depth biological and clinical phenotyping in the LBC1936 also enabled us to account for a broad range of vascular risk variables. A further strength of the LBC1936 is the availability of a measure of childhood cognitive ability. By including childhood cognitive ability as a covariate in our models, we were able eliminate its confounding effects on associations between SVD burden and later life cognitive abilities. Our study also has several limitations. Members of the LBC1936 are self-selecting, so represent a generally healthy, well-educated and highly-motivated sample and mostly have a mild non-clinical presentation of SVD. The main effect of this is probably a slight lowering of true effect sizes (61). It could be the case, therefore, that we are underestimating the associations between SVD burden and cognitive decline. However, that we observe associations between SVD burden and cognitive decline in a relatively healthy population of older individuals who are mostly free of overt cognitive impairment, demonstrates that SVD-related cognitive decline is present even before clinical presentation. A limitation of the longitudinal study design is that participants who dropped out of the study before Wave 5 may have done so due to poor health outcomes related to SVD (i.e. stroke or dementia).

Indeed, study non-completers had significantly greater baseline WMH volumes than participants who remained in the study up to Wave 5 (see Table S1). To some extent, we have been able to mitigate this survivor bias by using FIML as our model estimator, thus including all available data from our sample of 540 LBC1936 participants and ensuring that our results were not overly biased by the healthier participants of the initial 540, who completed all waves.

In this study we observed associations between the total MRI-visible burden of SVD and decline in general cognitive ability. The association we observed between SVD burden and decline in processing speed appears to be due to the overarching association between SVD burden and declining cognitive ability more generally. When monitoring SVD-related cognitive decline, trials of treatments or interventions for SVD should carry out an in-depth general cognitive ability (i.e. as opposed to a brief screening instrument) alongside assessments of any specific cognitive domain that is of particular interest. In doing so, any domain-specific cognitive changes can be examined in the context of declining general cognitive ability.

## Data Availability

Data supporting this study are available upon reasonable request from the corresponding authors.

## Conflict of Interest

None.

## Acknowledgements

We gratefully acknowledge contributions of the LBC1936 participants and members of the LBC research team who collect, handle and manage LBC data. The LBC1936 is supported by Age UK [MR/M01311/1] (http://www.disconnectedmind.ed.ac.uk) and the Medical Research Council [G1001245/96099]. LBC1936 MRI brain imaging was supported by Medical Research Council (MRC) grants [G0701120], [G1001245], [MR/M013111/1] and [MR/R024065/1]. OKLH is funded by the University of Edinburgh College of Medicine and Veterinary Medicine as part of the Wellcome Trust 4-year PhD in Translational Neuroscience at the University of Edinburgh. SRC, JMW, IJD, SMM and LB were supported by MRC grants [MR/M013111/1] and [MR/R024065/1]. SRC and IJD were additionally supported by a National Institutes of Health (NIH) research grant R01AG054628, and IJD was also supported by the Dementias Platform UK [MR/L015382/1]. JO was supported by the Economic and Social Research Council [ES/S015604/1]. MVH was supported by the Row Fogo Charitable Trust [BROD.FID3668413]. JMW is supported by the European Union Horizon 2020, (PHC-03-15, project no 666881), ‘SVDs@Target’, the Fondation Leducq Transatlantic Network of Excellence for the Study of Perivascular Spaces in Small Vessel Disease (ref no. 16 CVD 05), and the UK Dementia Research Institute which receives its funding from DRI Ltd, funded by the UK Medical Research Council, Alzheimer’s Society and Alzheimer’s Research UK.

## Notes

### Competing Interest Statement

The authors have declared no competing interest.

### Author Declarations

Approvals for Waves 2 to 5 of the LBC1936 were obtained from the Scotland A Research Ethics Committee for Scotland (07/MRE00/58).

