## Supplementary Tables S1-S6 for "Cerebral Small Vessel Disease Burden and Longitudinal Cognitive Decline from age 73 to 82: the Lothian Birth Cohort 1936"

*Table S1:* Baseline characteristics of study completers vs. participants lost to follow-up
*Table S2:* Unstandardised means and variances for the intercept and slope of each cognitive
*Table S3:* Results of factor-of-curves models of associations between total SVD burden and intercepts of latent cognitive variables
*Table S4:* Results of bifactor models of associations between total SVD burden and intercepts of latent cognitive variables
*Table S5:* Results of FoC models of associations between WMH volume/TIV and intercepts and slopes of latent cognitive variables between the ages of 73 and 82
*Table S6:* Fit indices for models presented in Table S5

*Table S1:* Baseline characteristics of study completers vs. participants lost to follow-up

| | n | Completers | n | Non-completers | *p*-value |
|---|---|---|---|---|---|
| **Sociodemographic** | | | | | |
| Age, years | 300 | 72.5 (0.7) | 240 | 72.5 (0.7) | 0.538 |
| Female, n (%) | 300 | 147 (49.0%) | 240 | 105 (43.8%) | 0.224 |
| Education, years | 300 | 11.0 (1.2) | 240 | 10.7 (1.1) | 0.014 |
| **Vascular risk** | | | | | |
| Hypertension history, n (%) | 300 | 135 (45.0%) | 240 | 124 (51.7%) | 0.123 |
| Systolic blood pressure | 297 | 145.6 (18.0) | 237 | 147.5 (18.1) | 0.229 |
| Diastolic blood pressure | 297 | 79.4 (9.2) | 237 | 80.1 (9.5) | 0.373 |
| Diabetes history, n (%) | 300 | 20 (6.7%) | 240 | 34 (14.2%) | 0.004 |
| HbA1c mmol/mol | 290 | 38.8 (5.8) | 228 | 39.3 (7.1) | 0.403 |
| Cholesterol, mmol/l | 291 | 5.3 (1.1) | 230 | 5.1 (1.2) | 0.043 |
| Cardiovascular disease history, n (%) | 300 | 83 (27.7%) | 240 | 71 (29.6%) | 0.624 |
| Smoking status, n (%) | 300 | Ever=141 (47.0%)<br>Never=159 (55.0%) | 240 | Ever=133 (55.4%)<br>Never=107 (44.6%) | 0.052 |
| **Cognitive** | | | | | |
| Moray House Test age 11 (max 76) | 283 | 51.2 (11.5) | 228 | 49.1 (12.3) | 0.046 |
| **Neuroimaging** | | | | | |
| Total WMH volume $cm^3$ | 298 | 10.47 (11.3) | 239 | 14.40 (14.3) | 0.004 |
| Total brain volume $cm^3$ | 300 | 991.9 (87.2) | 236 | 995.9 (90.1) | 0.612 |
| Lacunes, n (%) | 300 | Present=13 (4.3%)<br>Absent=287 (95.7%) | 240 | Present=15 (6.25%)<br>Absent=225 (93.75%) | 0.318 |
| Microbleeds, n (%) | 300 | Present=33 (11.0%)<br>Absent=267 (89.0%) | 240 | Present=32 (13.3%)<br>Absent=208 (86.7%) | 0.408 |

*Note:* Values are mean (standard deviation) unless otherwise specified. WMH: white matter hyperintensities of presumed vascular origin. Statistical comparisons performed using t-test for continuous variables and chi-squared test for binary variables. Total WMH volume was log transformed prior to statistical comparison due to right-sided skew.

*Table S2:* Unstandardised means and variances for the intercept and slope of each cognitive domain[a]

| | Intercepts | | Slopes | | | Fit indices | | | | |
|---|---|---|---|---|---|---|---|---|---|---|
| | Mean (SE) | Variance (SE) | Mean (SE) | Variance (SE) | SD change/year | $X^2$ | RMSEA | CFI | TLI | SRMR |
| **General cognitive ability** | 28.60 (0.26)*** | 18.91 (1.99)*** | -0.558 (0.03)*** | 0.140 (0.02)*** | -0.128 | 0.000 | 0.039 | 0.950 | 0.948 | 0.065 |
| **Processing speed** | 55.959 (0.25)*** | 7.990 (1.32)*** | -0.452 (0.04)*** | 0.140 (0.03)*** | -0.160 | 0.000 | 0.051 | 0.968 | 0.966 | 0.065 |
| **Memory** | 27.188 (0.41)*** | 42.821 (6.33)*** | -0.035 (0.05)*** | 0.540 (0.08)*** | -0.005 | 0.001 | 0.035 | 0.988 | 0.987 | 0.029 |
| **Visuospatial ability** | 17.365 (0.21)*** | 13.754 (1.47)*** | -0.303 (0.020)*** | 0.028 (0.013)* | -0.082 | 0.245 | 0.015 | 0.998 | 0.997 | 0.036 |

*Note:* Slopes represent change from age 73 to age 82. SD: standard deviation; SE: standard error. SD change per year calculated by dividing the slope mean by the intercept standard deviation. All p-values are uncorrected. * $p < 0.05$; ** $p < 0.01$; *** $p < 0.001$.

[a] Note that the cognitive domains will contain any variance due to general cognitive ability.

*Table S3:* Results of factor-of-curves models of associations between total SVD burden and intercepts of latent cognitive variables[a]

| | Intercept | | | |
|---|---|---|---|---|
| | **Standardised β (SE)** | **95% CI** | **Uncorrected p value** | **FDR-corrected p value[b]** |
| **General cognitive ability** | -0.377 (0.08) | -0.531, -0.224 | <0.001 | <0.001 |
| + age + sex | -0.346 (0.08) | -0.511, -0.182 | <0.001 | <0.001 |
| + age + sex + vascular risk | -0.334 (0.08) | -0.495, -0.174 | <0.001 | <0.001 |
| + age + sex + vascular risk + age-11 IQ | -0.299 (0.08) | -0.447, -0.151 | <0.001 | <0.001 |
| **Processing speed** | -0.402 (0.08) | -0.562, -0.243 | <0.001 | <0.001 |
| + age + sex | -0.364 (0.09) | -0.534, -0.193 | <0.001 | <0.001 |
| + age + sex + vascular risk | -0.351 (0.09) | -0.518, -0.183 | <0.001 | <0.001 |
| + age + sex + vascular risk + age-11 IQ | -0.322 (0.08) | -0.481, -0.164 | <0.001 | <0.001 |
| **Verbal memory** | -0.280 (0.09) | -0.447, -0.113 | 0.001 | 0.003 |
| + age + sex | -0.280 (0.08) | -0.441, -0.118 | 0.001 | 0.003 |
| + age + sex + vascular risk | -0.278 (0.08) | -0.440, -0.117 | 0.001 | 0.003 |
| + age + sex + vascular risk + age-11 IQ | -0.225 (0.08) | -0.383, -0.068 | 0.005 | 0.012 |
| **Visuospatial ability** | -0.240 (0.08) | -0.393, -0.086 | 0.002 | 0.005 |
| + age + sex | -0.220 (0.08) | -0.379, -0.062 | 0.007 | 0.016 |
| + age + sex + vascular risk | -0.209 (0.08) | -0.364, -0.055 | 0.008 | 0.017 |
| + age + sex + vascular risk + age-11 IQ | -0.173 (0.07) | -0.311, -0.036 | 0.014 | 0.022 |

*Note*: Four separate models were run for each cognitive factor, including covariates in a stepwise manner. Likelihood ratio test statistic (LR) and degrees of freedom (DF) for each of the unadjusted models were as follows: General cognitive ability (LR=6.79; DF=30), processing speed (LR=0.22; DF=2), verbal memory (LR=2.95; DF=1), visuospatial ability (LR=1.54; DF=2). CI: confidence interval; FDR: false discovery rate; SE: standard error. [a] Note that the cognitive domains will contain any variance due to general cognitive ability. [b]FDR correction was conducted across results presented in this table and in Table 3.

*Table S4:* Results of bifactor models of associations between total SVD burden and intercepts of latent cognitive variables

| | Intercept | | | |
|---|---|---|---|---|
| | Standardised β (SE) | 95% CI | Uncorrected p value | FDR-corrected p value[a] |
| **General cognitive ability** | -0.277 (0.13) | -0.528, -0.026 | 0.030 | 0.178 |
| + age + sex | -0.230 (0.11) | -0.452, -0.009 | 0.042 | 0.178 |
| + age + sex + vascular risk | -0.225 (0.12) | -0.450, 0.000 | 0.050 | 0.178 |
| + age + sex + vascular risk + age-11 IQ | -0.185 (0.11) | -0.408, 0.038 | 0.103 | 0.275 |
| **Processing speed** | -0.304 (0.15) | -0.603, -0.006 | 0.045 | 0.178 |
| + age + sex | -0.250 (0.13) | -0.498, -0.001 | 0.049 | 0.178 |
| + age + sex + vascular risk | -0.239 (0.13) | -0.489, 0.011 | 0.061 | 0.195 |
| + age + sex + vascular risk + age-11 IQ | -0.241 (0.13) | -0.501, 0.020 | 0.070 | 0.204 |
| **Verbal memory** | -0.115 (0.12) | -0.341, 0.111 | 0.318 | 0.678 |
| + age + sex | -0.111 (0.10) | -0.303, 0.081 | 0.258 | 0.590 |
| + age + sex + vascular risk | -0.119 (0.10) | -0.313, 0.075 | 0.231 | 0.569 |
| + age + sex + vascular risk + age-11 IQ | -0.093 (0.10) | -0.294, 0.108 | 0.366 | 0.732 |
| **Visuospatial ability** | 0.008 (0.14) | -0.256, 0.272 | 0.954 | 0.982 |
| + age + sex | -0.017 (0.16) | -0.320, 0.286 | 0.912 | 0.982 |
| + age + sex + vascular risk | -0.004 (0.16) | -0.306, 0.299 | 0.982 | 0.982 |
| + age + sex + vascular risk + age-11 IQ | 0.015 (0.16) | -0.293, 0.322 | 0.926 | 0.982 |

*Note*: Each bifactor model estimates associations between SVD burden and the four cognitive variables simultaneously. Four bifactor models, one without covariates and three further models, including covariates in a stepwise manner. Likelihood ratio test statistic (LR) and degrees of freedom (DF) for the unadjusted model was as follows: LR=55.3; DF=9. CI: confidence interval; FDR: false discovery rate; SE: standard error. [a]FDR correction was conducted across results presented in this table and in Table 4.

*Table S5:* Results of FoC models of associations between WMH volume/TIV and intercepts and slopes of latent cognitive variables between the ages of 73 and 82[a]

| | Intercept | | | | Slope | | | |
|---|---|---|---|---|---|---|---|---|
| | Standardised β (SE) | 95% CI | Uncorrected p value | FDR-corrected p value | Standardised β (SE) | 95% CI | Uncorrected p value | FDR-corrected p value |
| **General cognitive ability** | -0.226 (0.05) | -0.323, -0.129 | <0.001 | <0.001 | -0.127 (0.06) | -0.244, -0.009 | 0.035 | 0.047 |
| + age + sex | -0.222 (0.05) | -0.312, -0.133 | <0.001 | <0.001 | -0.149 (0.06) | -0.257, -0.040 | 0.007 | 0.012 |
| + age + sex + vascular risk | -0.216 (0.04) | -0.303, -0.130 | <0.001 | <0.001 | -0.148 (0.06) | -0.256, -0.039 | 0.008 | 0.012 |
| + age + sex + vascular risk + age-11 IQ | -0.194 (0.04) | -0.273, -0.115 | <0.001 | <0.001 | -0.149 (0.06) | -0.259, -0.039 | 0.008 | 0.012 |
| **Processing speed** | -0.231 (0.05) | -0.334, -0.128 | <0.001 | <0.001 | -0.148 (0.07) | -0.277, -0.018 | 0.026 | 0.036 |
| + age + sex | -0.230 (0.05) | -0.320, -0.140 | <0.001 | <0.001 | -0.177 (0.07) | -0.303, -0.050 | 0.006 | 0.012 |
| + age + sex + vascular risk | -0.224 (0.05) | -0.312, -0.136 | <0.001 | <0.001 | -0.176 (0.06) | -0.301, -0.050 | 0.006 | 0.012 |
| + age + sex + vascular risk + age-11 IQ | -0.205 (0.04) | -0.287, -0.123 | <0.001 | <0.001 | -0.176 (0.07) | -0.303, -0.049 | 0.007 | 0.012 |
| **Verbal memory** | -0.140 (0.05) | -0.235, -0.045 | 0.004 | 0.010 | -0.088 (0.08) | -0.236, 0.061 | 0.247 | 0.263 |
| + age + sex | -0.163 (0.05) | -0.251, -0.074 | <0.001 | <0.001 | -0.079 (0.07) | -0.220, 0.063 | 0.277 | 0.277 |
| + age + sex + vascular risk | -0.162 (0.05) | -0.251, -0.073 | <0.001 | <0.001 | -0.080 (0.07) | -0.220, 0.061 | 0.268 | 0.277 |
| + age + sex + vascular risk + age-11 IQ | -0.130 (0.04) | -0.216, -0.045 | 0.003 | 0.008 | -0.084 (0.07) | -0.226, 0.058 | 0.245 | 0.263 |
| **Visuospatial ability** | -0.161 (0.05) | -0.257, -0.066 | 0.001 | 0.003 | -0.178 (0.10) | -0.366, 0.011 | 0.064 | 0.073 |
| + age + sex | -0.136 (0.05) | -0.232, -0.039 | 0.006 | 0.012 | -0.197 (0.10) | -0.384, -0.010 | 0.039 | 0.048 |
| + age + sex + vascular risk | -0.130 (0.05) | -0.227, -0.033 | 0.008 | 0.012 | -0.198 (0.10) | -0.386, -0.011 | 0.038 | 0.048 |
| + age + sex + vascular risk + age-11 IQ | -0.111 (0.05) | -0.203, -0.020 | 0.017 | 0.025 | -0.190 (0.10) | -0.376, -0.004 | 0.046 | 0.055 |

*Note*: CI: confidence interval; FDR: false discovery rate; FoC: factor-of-curves; SE: standard error; TIV: total intracranial volume; WMH: white matter hyperintensities.
[a] Note that the cognitive domains will contain any variance due to general cognitive ability.

*Table S6:* Fit indices for models presented in Table S5

| | $X^2$ | RMSEA | CFI | TLI | SRMR |
|---|---|---|---|---|---|
| **General cognitive ability** | 0.000 | 0.039 | 0.944 | 0.942 | 0.063 |
| + age + sex | 0.000 | 0.037 | 0.941 | 0.937 | 0.068 |
| + age + sex + vascular risk | 0.000 | 0.036 | 0.941 | 0.937 | 0.067 |
| + age + sex + vascular risk + age-11 IQ | 0.000 | 0.037 | 0.938 | 0.933 | 0.066 |
| **Processing speed** | 0.000 | 0.045 | 0.967 | 0.964 | 0.064 |
| + age + sex | 0.000 | 0.034 | 0.972 | 0.969 | 0.060 |
| + age + sex + vascular risk | 0.000 | 0.033 | 0.972 | 0.969 | 0.060 |
| + age + sex + vascular risk + age-11 IQ | 0.000 | 0.034 | 0.969 | 0.966 | 0.059 |
| **Verbal memory** | 0.003 | 0.031 | 0.988 | 0.986 | 0.031 |
| + age + sex | 0.008 | 0.025 | 0.987 | 0.985 | 0.045 |
| + age + sex + vascular risk | 0.004 | 0.026 | 0.985 | 0.983 | 0.046 |
| + age + sex + vascular risk + age-11 IQ | 0.0017 | 0.026 | 0.983 | 0.981 | 0.047 |
| **Visuospatial ability** | 0.276 | 0.013 | 0.998 | 0.997 | 0.040 |
| + age + sex | 0.119 | 0.017 | 0.994 | 0.993 | 0.081 |
| + age + sex + vascular risk | 0.101 | 0.017 | 0.993 | 0.992 | 0.083 |
| + age + sex + vascular risk + age-11 IQ | 0.042 | 0.020 | 0.989 | 0.988 | 0.081 |

*Note:* CFI: comparative fit index; RMSEA: root mean square error of approximation; SRMR: standardized root mean square residual; TLI: Tucker Lewis index.
